# Children with tungiasis in Kenya have poor school performance and quality of life

**DOI:** 10.1101/2023.11.17.23298667

**Authors:** Lynne Elson, Christopher Kamau, Sammy Koech, Christopher Muthama, George Gichomba, Erastus Sinoti, Elwyn Chondo, Eliud Mburu, Miriam Wakio, Jimmy Lore, Marta Maia, Ifedayo Adetifa, Benedict Orindi, Phillip Bejon, Ulrike Fillinger

## Abstract

Tungiasis is a highly neglected tropical skin disease caused by the sand flea, *Tunga penetrans.* The flea burrows into the skin inducing a strong inflammatory response, leading to chronic pain and discomfort with potential impacts on quality of life. Few countries implement control efforts and there are few data on the impact of the disease to support policy decisions. We conducted a survey to determine the impact of tungiasis among primary school children across nine counties of Kenya.

A total of 10,600 pupils aged 8 to 14 years were randomly selected from 97 primary schools and examined for tungiasis. Those with tungiasis (83) were interviewed with respect to their quality of life using a modified dermatological quality of life index. For these cases and 576 randomly selected controls, school attendance and exam scores for maths, English and science were collected from school records. Mixed effect ordered logistic and linear models were used to assess associations between disease status and impact variables.

Compared to uninfected pupils, those with tungiasis missed more days of school (adjusted Incidence Rate Ratio (aIRR: 1.50, 95% CI: 1.03–2.21) and were less likely to receive a high score in maths (aOR 0.18, 95% CI: 0.08−0.40) and other subjects. Pupils with severe disease (>10 fleas) were five times more likely to experience severe pain and itching than those with mild disease (OR 5.0, 95% CI: 2.55−10.51) and a higher category of impact on their quality of life than those with mild disease (aOR 3.52, 95% CI: 1.22−10.17) when adjusted for covariates.

This study has demonstrated tungiasis has a considerable impact on children’s lives and academic achievement, equivalent to other diseases. This indicates the need for integrated disease management for school-aged children to protect their physical and cognitive development and their future prospects.

**Author Summary:** Tungiasis is one of the most neglected tropical diseases (NTDs). It is caused by a flea which burrows into the skin of the feet. The neglect has resulted in large gaps in our knowledge of the disease, how to treat and prevent infection. This is partly due to a lack of appreciation for the impact the disease has on patients’ lives by health officials, policymakers and the research community. In this study we aimed to determine the impact on school children in nine counties of Kenya. We found evidence that children experience considerable pain and itching which affects their ability to attend school and to concentrate on their classes. They had lower exam scores in maths, English and science than uninfected children. This study has provided evidence that tungiasis is a disease that affects children just as much as other diseases and urgently needs more attention from researchers, donors, and policy makers. Integrating tungiasis into comprehensive disease management for school-aged children will protect their physical and cognitive development and their future prospects.

## Introduction

Tungiasis is a highly neglected tropical skin disease which is widespread across sub-Saharan Africa[1]. In Kenya, human tungiasis is considered a significant individual and public health threat, with a national prevalence of 1.3% among children aged 8 to 14 years [2]. In selected communities, prevalence can reach as high as 64%, as in north-eastern Uganda [3–6]. Children under 15 years, elderly people and people with disabilities carry the highest disease burden [7].

Tungiasis is caused by female adult sand fleas (*Tunga penetrans*) which penetrate the skin of their mammalian hosts and stay embedded in one spot in the epidermis to mature and produce eggs [8]. The flea causes extensive morbidity resulting from an intense inflammatory response around the rapidly growing sand fleas [9]. The inflammation is further intensified by frequent bacterial superinfection and may result in tetanus, gangrene or septicemia[10]. The vast majority of the embedded sand fleas are located in the feet[1].

Little is known about the impact of tungiasis infection on a patient’s quality of life. The inflammation, pain and itching have been reported to affect children’s ability to sleep[9], walk[9, 11], attend school [12] and pay attention in class, thus reducing their school performance[12], but this has not been studied systematically. It was previously demonstrated in a small group of patients that tungiasis significantly impacts a child’s Quality of Life using the Dermatological Quality of Life Index (DLQI) modified to focus on a parasitic skin disease of the feet (TmDLQI) [13].

The objectives of this study were to determine the impact of tungiasis on children’s quality of life, on their school attendance and on their academic achievement in primary schools from nine counties of Kenya.

## Methods

### Study Population

This study was a cross-sectional survey embedded in a national level prevalence survey reported previously[2]. The survey was conducted in nine counties of Kenya, purposively selected to represent the five major climatic zones, cultures and geography of the country: Turkana, Samburu, Kericho, Nakuru, Muranga, Kajiado, Makueni, Taita Taveta and Kilifi. Within each county 22 primary schools were stratified randomly selected from pre-existing school list for each sub-county. The schools were surveyed in two phases, 11 schools per county in survey 1 from May to July 2021, and a different group of 11 schools in survey 2 from October 2021 to April 2023. Within each selected school an equal number of boys and girls were quasi-randomly selected by asking the pupils to assemble in three age groups; 8 and 9 years; 10 and 11 years; 12 to 14 years, and by sex within each age group. For each of these age/sex groups, every n^th^ pupil (n = total number in the group/ 19) was selected until 19 were selected, giving a total of 114 pupils overall.

All pupils were then asked to assent to a careful examination of their hands and feet for embedded *Tunga penetrans* fleas [2]. All infected pupils were also asked how much pain and how much itching they experienced in the past one week from the embedded fleas. They were asked to score this as either none, a little, some or a lot.

### Impact study sample size

The sample size for the impact assessments was based on a previous study on the impact of *Trichuris* infection on school absenteeism[14]. Children who were infected were absent from school for 28% of the school year, while uninfected children were absent for only 12% of the year. The sample size needed to detect this difference with 90% power and 95% confidence interval and a case: control ratio of 2, was 102 cases and 208 controls (Open Epi version 3). However, since we did not know if the impact would be as great as for trichuriasis, we applied a design effect of 2 for a total sample of 204 cases and 416 controls. This sample was spread across the nine counties and the 11 schools in each county, so that 3 cases and 6 controls were to be enrolled for each of 11 schools in each county to a total of 297 cases and 594 controls.

### Selection procedure

During survey 2, after the 114 pupils had been examined and their tungiasis infection status determined, six uninfected pupils were randomly selected using the lottery method. Every infected pupil identified was enrolled.

### Outcome variables

#### Anthropometrics

Measurements of pupils’ height and weight were taken using a tape measure and simple weighing scales, ensuring they were placed on a flat surface. From these height-for-age and weight-for-age z scores were calculated using the “zanthro” extension for Stata with the UK-WHO Term Growth Charts for adolescents provided[15].

#### School attendance and academic achievement

School attendance and exam records for the previous school term were provided by the teachers and searched for the selected pupils. Data were extracted on the number of days pupils were absent in the whole of the previous term as well as their exam results in math, English and science. The schools recorded exam scores differently for different school grades, a simple score of 0 to 4 for the lower grades, that is, one to four but an actual percentage mark for the upper grades, five to eight.

#### School grade delay

variable was created as follows. First, expected age in years for each school grade; grade 1 being 6.5, grade 2 being 7.5, grade 3 being 8.5 was created. Next, age difference was calculated for each pupil as a pupil’s age minus the expected age for his/her grade. This was then collapsed into a binary variable; 1 for any pupil who was more than 1.0 years older than the expected age for their grade and 0 for all others.

#### Pain and itching

All infected pupils were asked to score how much pain and how much itching they experienced in the past one week from the embedded fleas in their feet on a 4-point Likert scale of “none” (=0), “a little” (=1), “some” (=2), and “a lot” (=3).

#### Quality of Life

All infected pupils were interviewed following the Children’s Dermatological Quality of Life index (CDLQI)[16] previously modified for a skin disease of the feet, the Tungiasis modified Dermatological Quality of Life Index (TmDLQI) [13] and further modified with two additional questions added. Previous questions included: mobility, sleep, concentration in school, friendships, bullying and shame (six domains). We added feelings of anger and sadness to a total of eight domains. The instrument asked how much each of these was impacted by the embedded fleas in the past one week, scored on a 4-point Likert scale of “none” (=0), “a little” (=1), “some” (=2), and “a lot” (=3). An overall TmDLQI score was obtained by summing responses to all questions with a maximum of 24 for each pupil. As conducted with the previous mDLQI, the scores were then collapsed into quintiles (categories of impact) with thresholds based on a frequency histogram (S3, Supporting Information) and Waters et al [17]: 0–1 no impact, 2–5 small impact, 6–10 moderate, 11–15 large, 16–24 very large impact. The instrument was translated into Kiswahili. The tool was pre-tested and necessary adjustments made prior to the main survey.

### Explanatory variables

The main explanatory variable was disease severity assigned for each infected pupil as described previously[18] summing all the clinical signs into a clinical score and then dichotomised such that a pupil having a clinical score less than 11 being a mild case, and a pupil with a clinical score more than 10 being a severe case.

Other explanatory variables were county, school location (urban/ rural), school type (public/ private), pupil age, sex, disability, other skin abnormality, socioeconomic status (SES; as described previously[19]), adults the pupil lives with (both parents/ others), who the main caregiver is (mother/ other), mother’s school level attained (none/ primary/ secondary/ don’t know), father away a lot, mother away a lot, parents attend school meetings (never/sometimes/always), parents make sure pupils completes homework (never/sometimes/always), chronically ill family member, family member disabled, pupil missed school to help at home, pupil sleeps in parents’ house, number of people sleep in room with pupil.

## Data Analysis

All analyses were conducted in Stata IC version 15.1 (Stata Corp LLC, College Station, Texas, USA). First, we described the participants by their background characteristics and disease status. Next, univariable mixed effect models were fitted for outcome and all the explanatory variables. Height-for-age and weight-for-age were also included as covariates in models for the other five outcomes. Only covariates with a p-value below 0.200 were included in the multivariable models and stepwise backward elimination used to reach the final models with the lowest Akaike Information Criteria (AIC). Significance of categorical variables in the final model were confirmed using Wald Chi tests. Results for the final models are presented in the main body of the manuscript while univariable analyses are presented in the supplementary material, additional file 2. School was included as a random effect in all models. Depending on the nature of the outcome, we fitted either a linear mixed effects model (anthropometrics, academic achievement for upper grade), a negative binomial mixed effects model (absenteeism), a logit mixed effects model (school grade delay) or ordered logit mixed effects model (academic achievement for lower grade, pain and itching, and TmDLQI quintiles).

To identify the quality-of-life domain most affected by tungiasis the percent of infected pupils scoring a 2 or 3 for each domain was calculated. A two-level ordered logistic regression analysis with school as a random effect was used to test for association of each domain with disease severity. We performed these analyses under the valid missing at random assumption, as we used likelihood approaches [20].

## Results

### Study Population

A total of 659 pupils from nine counties were enrolled and analysed. The majority (585) of these pupils were from public schools and rural areas (577). The median age was 11 years (IQR 9−13) and 50.9% were boys. Only 83 (12.6%) pupils had tungiasis, 10 (1.5%) were disabled and 42 (6.4%) had some other skin abnormality. There were no pupils with tungiasis in the private schools (Additional file 1 in supplementary material).

### Anthropometrics

Of the 659 pupils, 117 (17.75%) were classified as underweight (weight-for-age z-score<-2.0) while 47 (7.13%) were stunted (height-for-age z-score<-2.0). Of the 83 infected pupils, 32 (38.55%) were underweight and 10 (12.05%) were stunted (See supplementary table 1). Univariable mixed effect linear models for weight-for-age found a significant negative association with tungiasis status (β - 0.65, 95% *CI:* -0.99−-0.31). This negative association remained (β -0.41, 95% *CI:* -0.75−-0.06) even when adjusted for other covariables, including SES, and whether a child lived with both parents (Table 1). There was no association of tungiasis with height-for-age (β -0.09, 95% CI: -0.47−0.28).

**Table 1.**
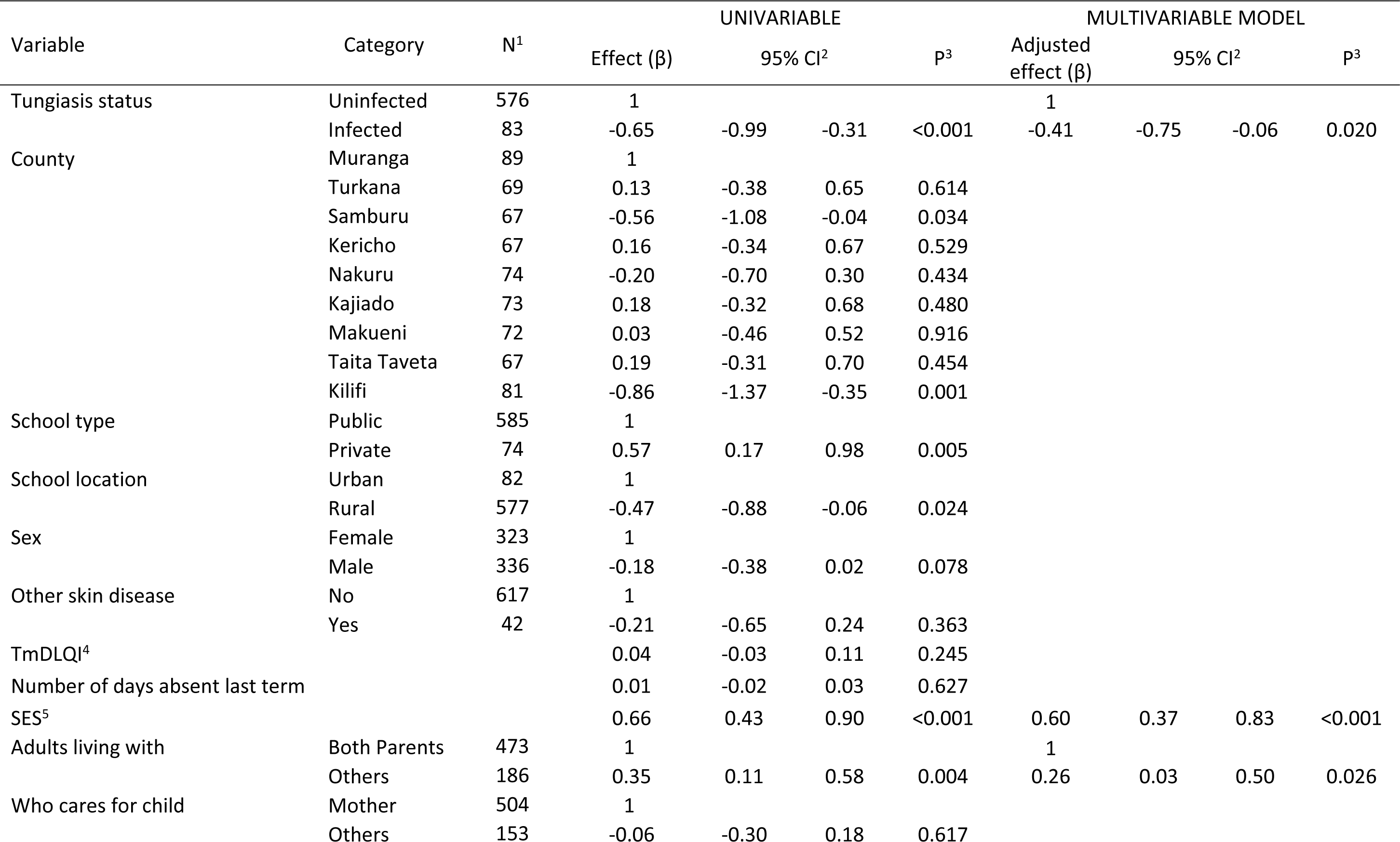
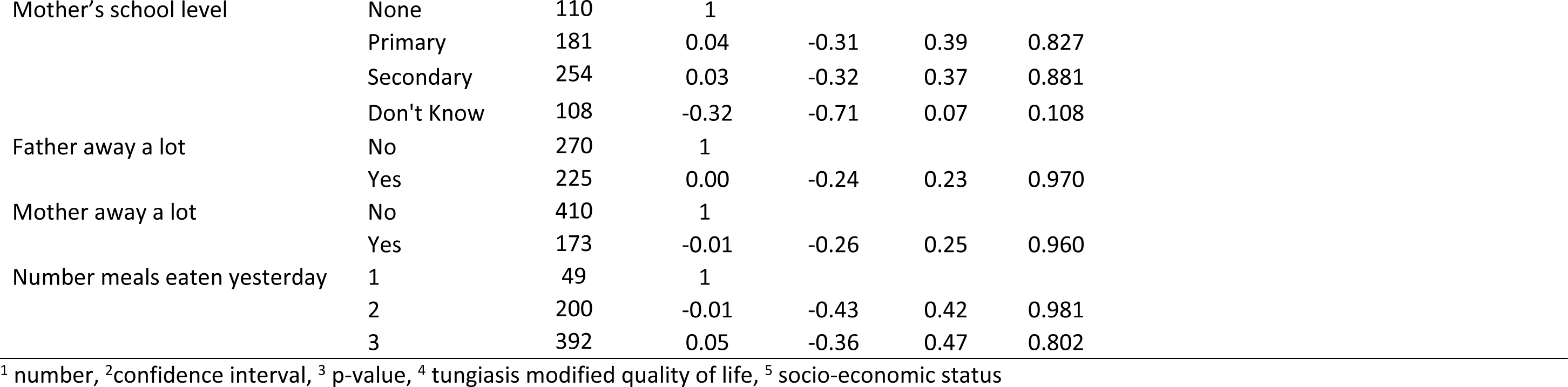
Linear mixed effects regression analysis for nutrition status (weight-for-age z-scores)

| Variable | Category | N<sup>1</sup> | UNIVARIABLE |  |  |  | MULTIVARIABLE MODEL |  |  |  |
| --- | --- | --- | --- | --- | --- | --- | --- | --- | --- | --- |
| | | | Effect ($\beta$) | 95% CI<sup>2</sup> | | P<sup>3</sup> | Adjusted effect ($\beta$) | 95% CI<sup>2</sup> | | P<sup>3</sup> |
| Tungiasis status | Uninfected | 576 | 1 |  |  |  | 1 |  |  |  |
|  | Infected | 83 | -0.65 | -0.99 | -0.31 | <0.001 | -0.41 | -0.75 | -0.06 | 0.020 |
| County | Muranga | 89 | 1 |  |  |  |  |  |  |  |
|  | Turkana | 69 | 0.13 | -0.38 | 0.65 | 0.614 |  |  |  |  |
|  | Samburu | 67 | -0.56 | -1.08 | -0.04 | 0.034 |  |  |  |  |
|  | Kericho | 67 | 0.16 | -0.34 | 0.67 | 0.529 |  |  |  |  |
|  | Nakuru | 74 | -0.20 | -0.70 | 0.30 | 0.434 |  |  |  |  |
|  | Kajiado | 73 | 0.18 | -0.32 | 0.68 | 0.480 |  |  |  |  |
|  | Makueni | 72 | 0.03 | -0.46 | 0.52 | 0.916 |  |  |  |  |
|  | Taita Taveta | 67 | 0.19 | -0.31 | 0.70 | 0.454 |  |  |  |  |
|  | Kilifi | 81 | -0.86 | -1.37 | -0.35 | 0.001 |  |  |  |  |
| School type | Public | 585 | 1 |  |  |  |  |  |  |  |
|  | Private | 74 | 0.57 | 0.17 | 0.98 | 0.005 |  |  |  |  |
| School location | Urban | 82 | 1 |  |  |  |  |  |  |  |
|  | Rural | 577 | -0.47 | -0.88 | -0.06 | 0.024 |  |  |  |  |
| Sex | Female | 323 | 1 |  |  |  |  |  |  |  |
|  | Male | 336 | -0.18 | -0.38 | 0.02 | 0.078 |  |  |  |  |
| Other skin disease | No | 617 | 1 |  |  |  |  |  |  |  |
|  | Yes | 42 | -0.21 | -0.65 | 0.24 | 0.363 |  |  |  |  |
| TmDLQI<sup>4</sup> |  |  | 0.04 | -0.03 | 0.11 | 0.245 |  |  |  |  |
| Number of days absent last term |  |  | 0.01 | -0.02 | 0.03 | 0.627 |  |  |  |  |
| SES<sup>5</sup> |  |  | 0.66 | 0.43 | 0.90 | <0.001 | 0.60 | 0.37 | 0.83 | <0.001 |
| Adults living with | Both Parents | 473 | 1 |  |  |  | 1 |  |  |  |
|  | Others | 186 | 0.35 | 0.11 | 0.58 | 0.004 | 0.26 | 0.03 | 0.50 | 0.026 |
| Who cares for child | Mother | 504 | 1 |  |  |  |  |  |  |  |
|  | Others | 153 | -0.06 | -0.30 | 0.18 | 0.617 |  |  |  |  |

|  |  |  |  |  |  |  |
| --- | --- | --- | --- | --- | --- | --- |
| Mother's school level | None | 110 | 1 |  |  |  |
|  | Primary | 181 | 0.04 | -0.31 | 0.39 | 0.827 |
|  | Secondary | 254 | 0.03 | -0.32 | 0.37 | 0.881 |
|  | Don't Know | 108 | -0.32 | -0.71 | 0.07 | 0.108 |
| Father away a lot | No | 270 | 1 |  |  |  |
|  | Yes | 225 | 0.00 | -0.24 | 0.23 | 0.970 |
| Mother away a lot | No | 410 | 1 |  |  |  |
|  | Yes | 173 | -0.01 | -0.26 | 0.25 | 0.960 |
| Number meals eaten yesterday | 1 | 49 | 1 |  |  |  |
|  | 2 | 200 | -0.01 | -0.43 | 0.42 | 0.981 |
|  | 3 | 392 | 0.05 | -0.36 | 0.47 | 0.802 |

<sup>1</sup> number, <sup>2</sup>confidence interval, <sup>3</sup> p-value, <sup>4</sup> tungiasis modified quality of life, <sup>5</sup> socio-economic status

### Absenteeism

Records of the number of days a pupil was absent during the previous school term were recorded for 624 pupils, of whom 74 were infected. Univariable analyses found pupils with tungiasis missed nearly twice as many days of school than uninfected pupils (IRR: 1.87, 95% *CI*: 1.27–2.75), but many other covariates were also associated with absenteeism, that could account for this difference (Table 2). When combined in a multivariable model, tungiasis infection was still associated, with infected pupils missing 1.5 times more days than uninfected pupils (aIRR: 1.50, 95% CI: 1.03–2.21) even when adjusted for county, age, SES, caregiver school level achieved and family disability (Table 2).

**Table 2.**
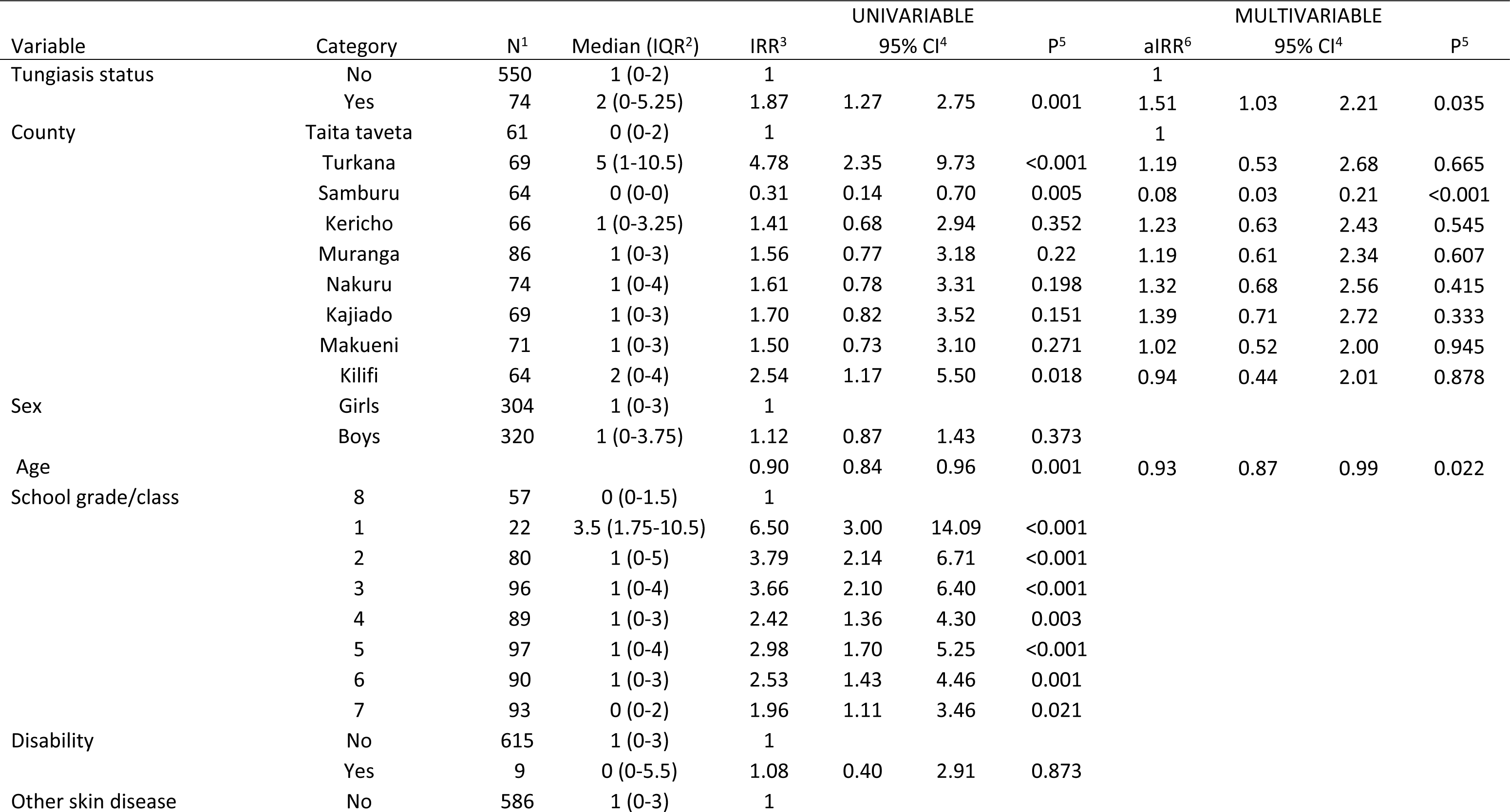
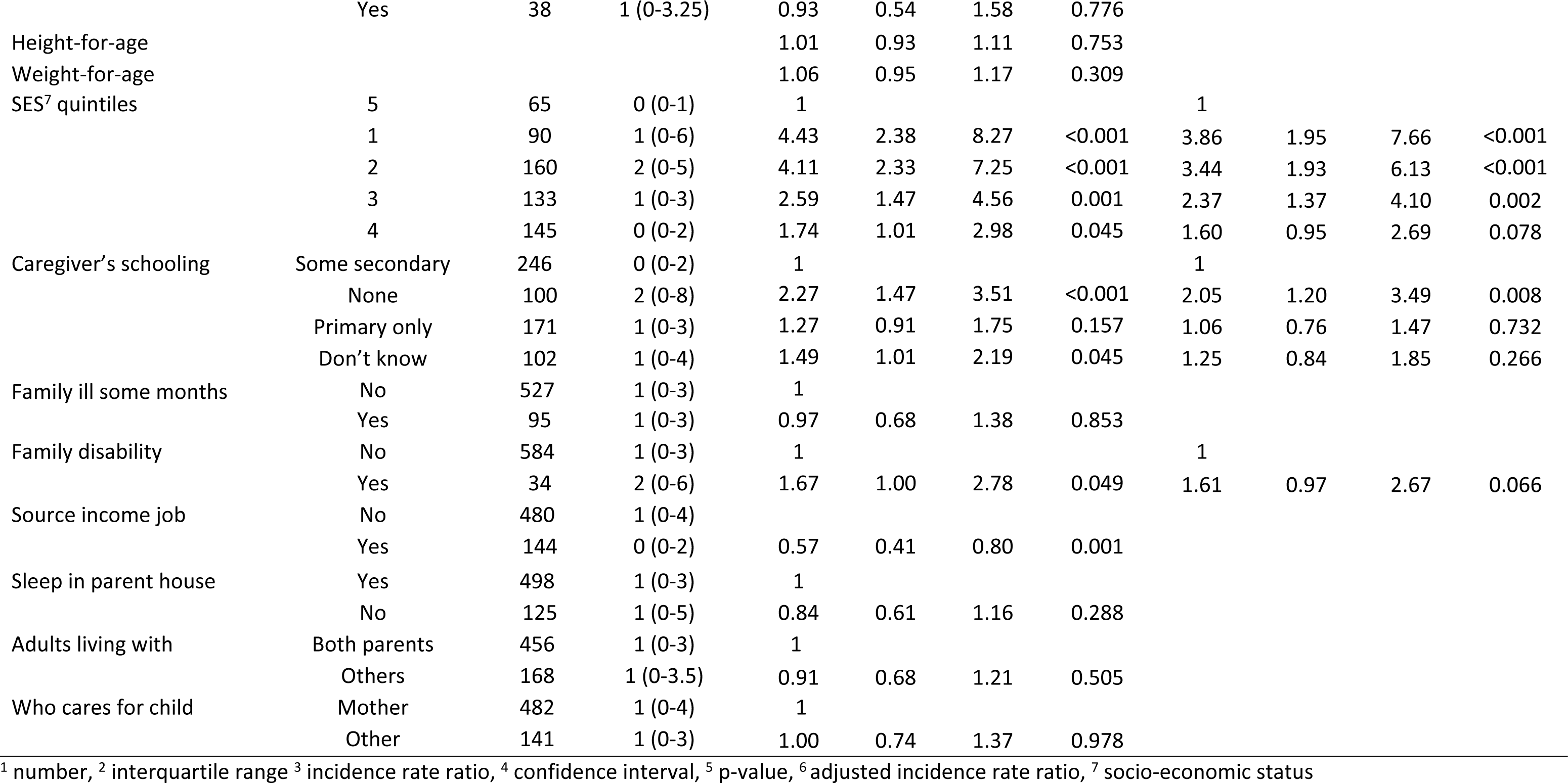
Negative binomial regression for school absenteeism (number of days absent).

### School performance

Of the 659 pupils, exam results were obtained for 619 of whom 273 were from the lower grades with a median age of 9 years (IQR 8-10.5 years), for whom results were scored on a Likert-type scale of 0 to 4. Another 346 pupils were from the upper grades with a median age of 12 years (IQR 11-14 years) for whom actual percentage exam results were obtained.

For the lower classes the regression models indicated pupils with tungiasis had five times lower odds of achieving the higher scores compared to uninfected pupils (maths aOR 0.18, 95% CI: 0.08−0.40, English aOR 0.19, 95% CI: 0.08−0.44, and science aOR 0.20, 95% CI: 0.09−0.44), even when adjusted for age, disability, the number of days absent from school in the past term, SES, whether the mother was away a lot and the height-for-age z-scores (Table 3). There was no association of school type (public or private) nor weight-for-age z-scores with any subject exam results in this age group.

**Table 3.**
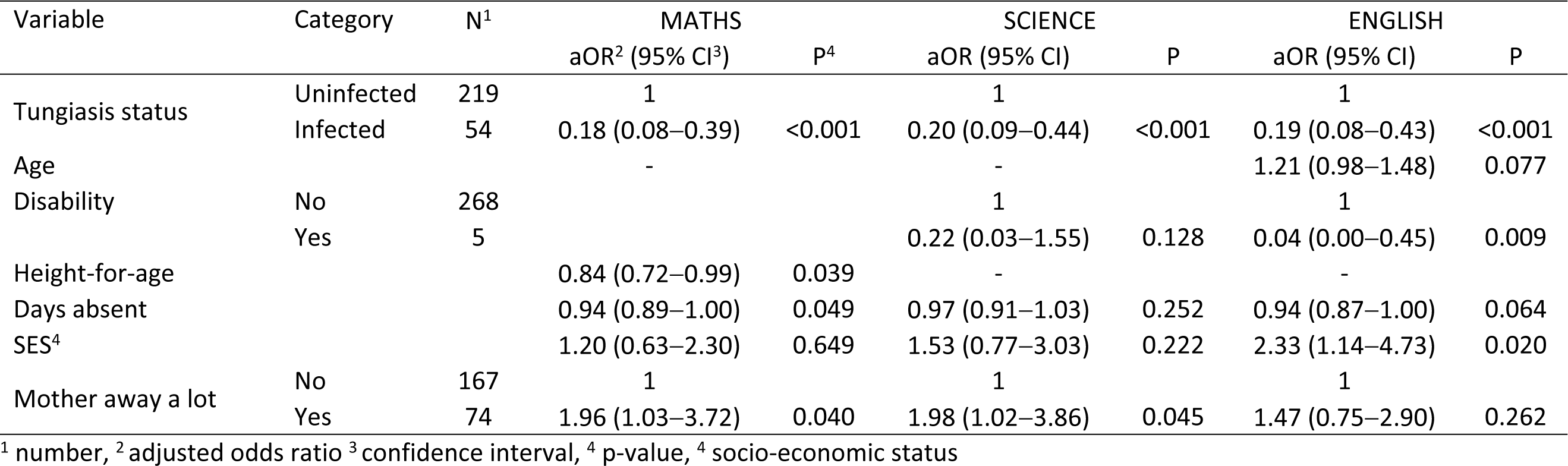
Panel Ordered Logistic Regression analysis of school exam results for Maths, Science and English for pupils in grade 1 to 4. (Univariable tables in Supporting Information S2)

| Variable | Category | N<sup>1</sup> | MATHS |  | SCIENCE |  | ENGLISH |  |
| --- | --- | --- | --- | --- | --- | --- | --- | --- |
|  |  |  | aOR<sup>2</sup> (95% CI<sup>3</sup>) | P<sup>4</sup> | aOR (95% CI) | P | aOR (95% CI) | P |
| Tungiasis status | Uninfected | 219 | 1 |  | 1 |  | 1 |  |
|  | Infected | 54 | 0.18 (0.08–0.39) | <0.001 | 0.20 (0.09–0.44) | <0.001 | 0.19 (0.08–0.43) | <0.001 |
| Age |  |  | - |  | - |  | 1.21 (0.98–1.48) | 0.077 |
| Disability | No | 268 |  |  | 1 |  | 1 |  |
|  | Yes | 5 |  |  | 0.22 (0.03–1.55) | 0.128 | 0.04 (0.00–0.45) | 0.009 |
| Height-for-age |  |  | 0.84 (0.72–0.99) | 0.039 | - |  | - |  |
| Days absent |  |  | 0.94 (0.89–1.00) | 0.049 | 0.97 (0.91–1.03) | 0.252 | 0.94 (0.87–1.00) | 0.064 |
| SES<sup>4</sup> |  |  | 1.20 (0.63–2.30) | 0.649 | 1.53 (0.77–3.03) | 0.222 | 2.33 (1.14–4.73) | 0.020 |
| Mother away a lot | No | 167 | 1 |  | 1 |  | 1 |  |
|  | Yes | 74 | 1.96 (1.03–3.72) | 0.040 | 1.98 (1.02–3.86) | 0.045 | 1.47 (0.75–2.90) | 0.262 |

<sup>1</sup> number, <sup>2</sup> adjusted odds ratio <sup>3</sup> confidence interval, <sup>4</sup> p-value, <sup>4</sup> socio-economic status

The association of school performance with tungiasis was not found for the older pupils in the upper grades, the other independent variables being more strongly associated (Table 4). For each exam subject a different set of variables were associated, but common to all and most strongly associated for all subjects was attending a private school rather than a public school. Repeating the analyses for pupils in the public schools only, did not change the lack of association of tungiasis with exam results for the older pupils (data not shown).

**Table 4.** Linear Regression analysis of school exam results in Maths, Science and English for pupils in grade 5 to 8. (see Univariable tables in Supporting Information S2).

| Variable | Category | N<sup>1</sup> | MATHS<br>β (95% CI)<sup>2</sup> | P<sup>3</sup> | SCIENCE<br>β (95% CI) | P | ENGLISH<br>β (95% CI) | P |
| --- | --- | --- | --- | --- | --- | --- | --- | --- |
| Tungiasis status | Uninfected | 326 | 1 |  | 1 |  | 1 |  |
|  | Infected | 20 | -8.52 (-17.03– -0.02) | 0.050 | -6.03 (-15.45–3.40) | 0.210 | -4.57 (-12.20–3.07) | 0.241 |
| School type | Public | 301 | 1 |  | 1 |  | 1 |  |
|  | Private | 45 | 15.39 (7.73–23.04) | <0.001 | 18.06 (8.82–27.30) | <0.001 | 8.96 (3.17–14.74) | 0.002 |
| School location | Urban | 45 |  |  | 1 |  |  |  |
|  | Rural | 301 |  |  | 8.81 (-0.22–17.84) | 0.056 |  |  |
| Age |  |  |  |  |  |  | -2.72 (-3.91– -1.54) | <0.001 |
| Weight-for-age |  |  |  |  |  |  | -1.18 (-2.74–0.38) | 0.138 |
| Days absent |  |  |  |  | -0.49 (-1.14–0.16) | 0.142 | -0.51 (-1.10–0.08) | 0.091 |
| SES<sup>4</sup> |  |  | 4.07 (-1.03–9.16) | 0.117 | 3.80 (-2.10–9.70) | 0.206 | 7.47 (2.44–12.50) | 0.004 |
| Adults living with | Both parents | 249 |  |  |  |  | 1 |  |
|  | Other | 97 |  |  |  |  | 4.04 (0.25–7.84) | 0.037 |
| Mother schooling level | None | 65 | 1 |  | 1 |  | 1 |  |
|  | Primary | 98 | -5.38 (-11.37–0.61) | 0.078 | -5.74 (-12.44–0.96) | 0.093 | -0.69 (-6.37–4.98) | 0.811 |
|  | Secondary | 138 | -1.46 (-7.90–4.99) | 0.658 | 0.18 (-7.23–7.59) | 0.962 | 1.39 (-4.71–7.49) | 0.655 |
|  | Don't know | 42 | -1.20 (-8.08–5.67) | 0.732 | 2.05 (-5.97–10.07) | 0.616 | 0.71 (-5.80–7.22) | 0.831 |
| Father away a lot | No | 145 |  |  | 1 |  |  |  |
|  | Yes | 118 |  |  | 2.08 (-1.87–6.03) | 0.302 |  |  |
| Parents attend school meetings | Never | 14 | 1 |  |  |  |  |  |
|  | Sometimes | 137 | 2.11 (-7.57–11.79) | 0.669 |  |  |  |  |
|  | Always | 194 | 4.41 (-5.50–14.31) | 0.383 |  |  |  |  |
| Parent make sure do homework | Never | 40 | 1 |  | 1 |  | 1 |  |
|  | Sometimes | 122 | 1.23 (-5.08–7.54) | 0.702 | 3.43 (-3.87–10.74) | 0.357 | 3.85 (-2.18–9.88) | 0.211 |
|  | Always | 183 | 2.19 (-4.52–8.91) | 0.522 | 8.92 (1.62–16.23 | 0.017 | 6.90 (0.76–13.03) | 0.028 |
| Family member ill some months | No | 282 | 1 |  |  |  | 1 |  |
|  | Yes | 62 | -3.58 (-8.08–0.93) | 0.119 |  |  | -2.05 (-6.60–2.51) | 0.378 |
| Miss school to help parents | No | 288 | 1 |  | 1 |  | 1 |  |
|  | Yes | 53 | -4.02 (-9.17–1.13) | 0.126 | -1.25 (-7.32–4.81) | 0.685 | -3.11 (-8.41–2.20) | 0.251 |
| Sleep in parent house | No | 81 |  |  | 1 |  |  |  |
|  | Yes | 265 |  |  | 4.99 (0.31–9.68) | 0.037 |  |  |

<sup>1</sup> number, <sup>2</sup> confidence interval, <sup>3</sup> p-value, <sup>4</sup> socio-economic status

### Delay in school grade

Of the 659 pupils enrolled, 253 (38.4%) were more than one year older than expected for their school grade and of these 47 (18.6%) were infected, compared to only 36 of the 406 pupils (8.9%) who were the expected age. While school grade delay was associated with tungiasis in the univariable analyses (OR 2.10, 95% *CI*: 1.12−3.94), several other independent covariables were too, and it was these variables that were more strongly associated with school grade delay and not tungiasis (aOR 1.27, 95% *CI*: 0.58−2.78) (Table 5).

**Table 5.** Logistic regression analysis for delay in school grade (delayed at least one year):

| Variable | Category | N<sup>1</sup> | OR<sup>2</sup> | UNIVARIABLE |  |  | MULTIVARIABLE |  |  |  |
| --- | --- | --- | --- | --- | --- | --- | --- | --- | --- | --- |
|  |  |  |  | 95% CI<sup>3</sup> |  | P<sup>4</sup> | aOR<sup>5</sup> | 95% CI |  | P |
| Tungiasis status | Not infected | 576 | 1 |  |  |  | 1 |  |  |  |
|  | Infected | 83 | 2.10 | 1.12 | 3.94 | 0.022 | 1.27 | 0.58 | 2.78 | 0.549 |
| County | Muranga | 89 | 1 |  |  |  | 1 |  |  |  |
|  | Turkana | 69 | 11.44 | 4.21 | 31.07 | <0.001 | 4.74 | 1.10 | 20.40 | 0.037 |
|  | Samburu | 67 | 3.76 | 1.40 | 10.11 | 0.009 | 2.23 | 0.60 | 8.24 | 0.229 |
|  | Kericho | 67 | 2.09 | 0.76 | 5.70 | 0.152 | 1.72 | 0.52 | 5.73 | 0.378 |
|  | Nakuru | 74 | 1.64 | 0.60 | 4.51 | 0.338 | 1.17 | 0.35 | 3.91 | 0.802 |
|  | Kajiado | 73 | 2.07 | 0.76 | 5.60 | 0.153 | 1.91 | 0.56 | 6.49 | 0.300 |
|  | Makueni | 72 | 1.46 | 0.54 | 3.96 | 0.453 | 0.88 | 0.26 | 2.98 | 0.833 |
|  | Taita Taveta | 67 | 1.29 | 0.46 | 3.63 | 0.636 | 0.83 | 0.24 | 2.90 | 0.766 |
|  | Kilifi | 81 | 20.42 | 7.12 | 58.53 | <0.001 | 12.67 | 3.01 | 53.32 | 0.001 |
| School type | Public | 585 | 1 |  |  |  |  |  |  |  |
|  | Private | 74 | 0.15 | 0.05 | 0.44 | 0.001 |  |  |  |  |
| School location | Urban | 82 | 1 |  |  |  |  |  |  |  |
|  | Rural | 577 | 3.19 | 1.20 | 8.50 | 0.020 |  |  |  |  |
| Age |  |  | 1.39 | 1.24 | 1.56 | <0.001 | 1.49 | 1.30 | 1.71 | <0.001 |
| Sex | Female | 323 | 1 |  |  |  |  |  |  |  |
|  | Male | 336 | 1.47 | 1.00 | 2.16 | 0.049 |  |  |  |  |
| Disability | No | 649 | 1 |  |  |  |  |  |  |  |
|  | Yes | 10 | 2.44 | 0.46 | 12.90 | 0.294 |  |  |  |  |
| Other skin abnormality | No | 617 | 1 |  |  |  |  |  |  |  |
|  | Yes | 42 | 1.44 | 0.64 | 3.26 | 0.377 |  |  |  |  |
| Height-for-age |  |  | 0.70 | 0.60 | 0.82 | <0.001 |  |  |  |  |
| Weight-for-age |  |  | 0.77 | 0.67 | 0.89 | <0.001 | 0.72 | 0.59 | 0.87 | 0.001 |
| Days absent |  |  | 1.10 | 1.04 | 1.17 | 0.001 | 1.11 | 1.04 | 1.19 | 0.002 |
| SES<sup>6</sup> |  |  | 0.22 | 0.14 | 0.35 | <0.001 | 0.42 | 0.21 | 0.83 | 0.013 |
| Adults living with | Both parents | 473 | 1 |  |  |  |  |  |  |  |
|  | Other adults | 186 | 1.14 | 0.74 | 1.76 | 0.543 |  |  |  |  |
| Who cares for child | Mother | 504 | 1 |  |  |  |  |  |  |  |
|  | Others | 153 | 0.75 | 0.47 | 1.19 | 0.219 |  |  |  |  |
| Mother's schooling | None | 110 | 1 |  |  |  | 1 |  |  |  |
|  | Primary | 181 | 0.69 | 0.36 | 1.33 | 0.264 | 2.15 | 0.87 | 5.28 | 0.095 |
|  | Secondary | 254 | 0.56 | 0.28 | 1.10 | 0.092 | 2.63 | 1.01 | 6.84 | 0.048 |
|  | Don't know | 108 | 0.69 | 0.33 | 1.47 | 0.341 | 2.26 | 0.80 | 6.35 | 0.122 |
| Father away a lot | No | 270 | 1 |  |  |  |  |  |  |  |
|  | Yes | 228 | 0.96 | 0.61 | 1.52 | 0.875 |  |  |  |  |
| Mother away a lot | No | 410 | 1 |  |  |  |  |  |  |  |
|  | Yes | 173 | 0.99 | 0.60 | 1.64 | 0.972 |  |  |  |  |
| Parents attend school meetings | Never | 38 | 1 |  |  |  |  |  |  |  |
|  | Sometimes | 260 | 1.50 | 0.66 | 3.42 | 0.338 |  |  |  |  |
|  | Always | 358 | 0.89 | 0.38 | 2.06 | 0.781 |  |  |  |  |
| Parents check homework done | Never | 95 | 1 |  |  |  |  |  |  |  |
|  | Sometimes | 235 | 0.94 | 0.52 | 1.70 | 0.834 |  |  |  |  |
|  | Always | 328 | 0.44 | 0.24 | 0.81 | 0.008 |  |  |  |  |
| Family member ill some months | No | 548 | 1 |  |  |  |  |  |  |  |
|  | Yes | 108 | 1.06 | 0.63 | 1.78 | 0.824 |  |  |  |  |
| Family member has disability | No | 616 | 1 |  |  |  |  |  |  |  |
|  | Yes | 37 | 1.52 | 0.66 | 3.49 | 0.326 |  |  |  |  |
| Miss school to help at home | No | 548 | 1 |  |  |  | 1 |  |  |  |
|  | Yes | 102 | 2.14 | 1.25 | 3.65 | 0.005 | 1.65 | 0.86 | 3.17 | 0.134 |
| Sleep in parents' house | No | 133 | 1 |  |  |  |  |  |  |  |
|  | Yes | 524 | 0.64 | 0.39 | 1.04 | 0.072 |  |  |  |  |

<sup>1</sup> number, <sup>2</sup> odds ratio <sup>3</sup> confidence interval, <sup>4</sup> p-value, <sup>5</sup> adjusted odds ratio <sup>6</sup> socio-economic status

### Pain and Itching

All 230 infected pupils identified during both survey 1 and 2 were asked about the amount of pain they experienced from the embedded fleas and the amount of itching the fleas caused them in the previous one week, scored from 0 (= none) to 3 (= a lot). Univariable models demonstrated that pupils with severe tungiasis (more than 10 fleas) were five times more likely to experience higher levels of pain than the pupils with mild tungiasis (OR 4.99, 95% CI 2.50−9.99). The same was true for itching (OR 5.44, 95% CI 2.65−11.19).

### Quality of Life

The domains of the Tungiasis-modified Dermatological Life Quality Index (TmDLQI) with the most pupils expressing “some” or “a lot” of impact were shame, disturbed sleep and difficulty concentrating at school due to the itching (Table 6). Using panel ordered univariable models it was clear mild cases felt the same levels of shame as severe cases. However, pupils with severe tungiasis had 4 times higher odds of experiencing higher levels of impact on concentration and mobility (Table 6). Although the p-values were not less than 0.05, pupils with severe disease had 3 times higher odds of experiencing higher levels of bullying and disturbed sleep.

**Table 6.** Association of Tungiasis associated-quality-of-life domains for pupils with severe disease compared to pupils with mild disease.

| Quality of Life domain | % scoring “some”<br>or “a lot” of impact | Odds<br>Ratio | 95% Conf.<br>Interval |  | P-value |
| --- | --- | --- | --- | --- | --- |
| Shame | 41.8 | 1.15 | 0.31 | 4.20 | 0.836 |
| Disturbed sleep | 35.8 | 2.70 | 0.96 | 7.54 | 0.059 |
| Difficulty concentrating due to itch | 32.8 | 4.19 | 1.06 | 16.48 | 0.041 |
| Feel anger | 31.3 | 1.25 | 0.42 | 3.75 | 0.690 |
| Mobility | 25.4 | 4.15 | 1.29 | 13.36 | 0.017 |
| Bullying | 23.9 | 3.43 | 0.94 | 12.54 | 0.062 |
| Affects friendship | 28.4 | 0.79 | 0.27 | 2.34 | 0.674 |
| Feel sad | 28.4 | 1.37 | 0.42 | 4.47 | 0.602 |

When all of the domain scores were summed for each pupil to obtain the TmDLQI score, the median score was 9 (IQR 4–13) and ranged from 0 to 24 (the maximum possible). When collapsed into quintiles, 80% of severe cases reported a moderate to very large impact on quality of life (TmDLQI 6–24), while only 62.5% of mild cases were considered to have this level of impact (Figure 1). Since quality of life could be impacted by many factors in a child’s life, we conducted multivariable analysis with the same set of explanatory variables as used above with the TmDLQI quintiles and found pupils with severe tungiasis had a nearly four times higher odds of experiencing higher category of impact than those with mild tungiasis (aOR 3.52, 95% *CI*: 1.22–10.17) even when adjusted for age, and the mother being away a lot (Table 7). The older the child the less the impact, but children whose mothers were away a lot had 3 times higher odds of experiencing a higher category of impact.

**Figure 1.**
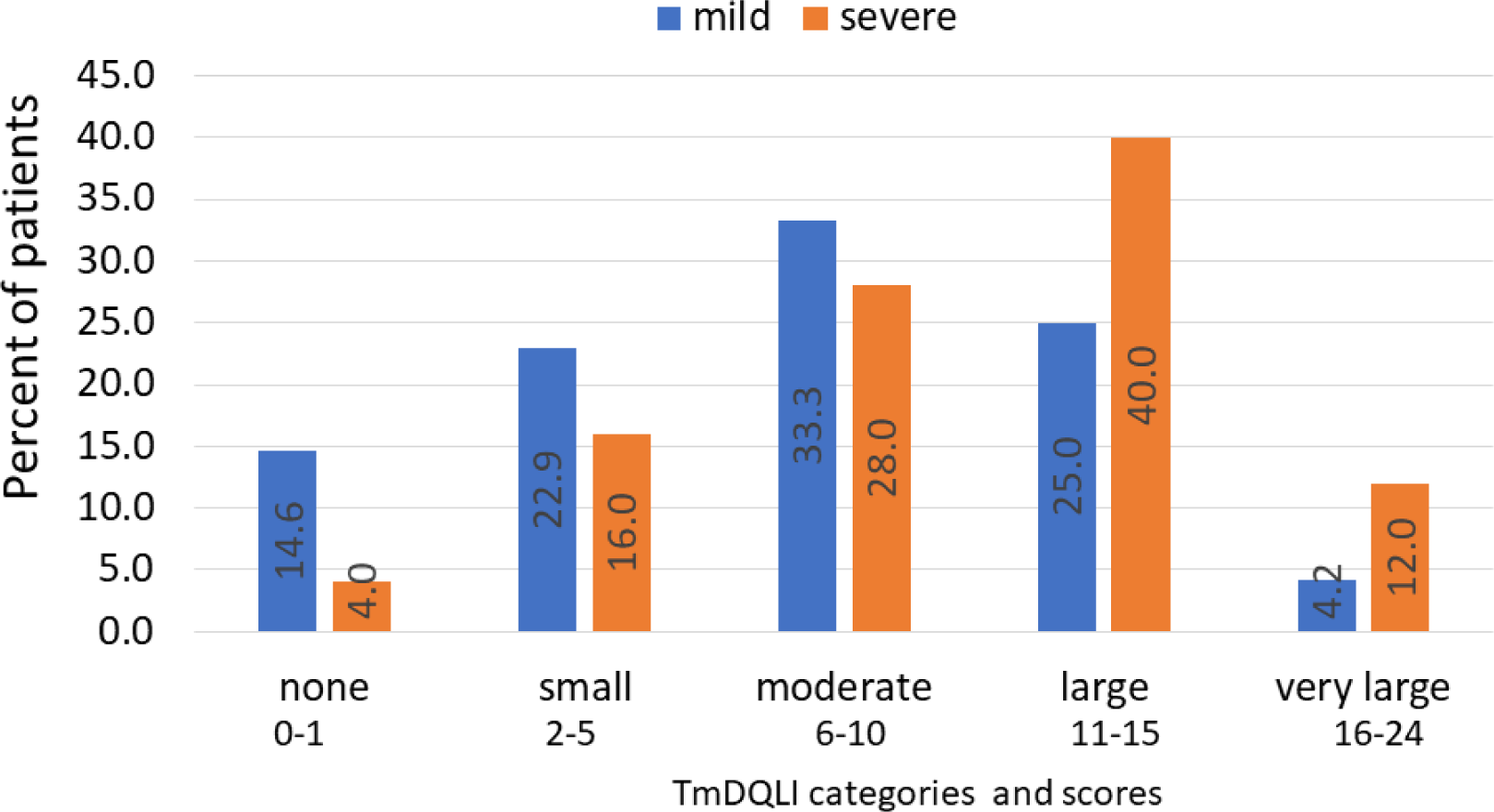
Distribution of tungiasis-associated quality-of-life categories by disease severity. Severe disease: orange bars, mild disease: blue bars.

**Table 7.**
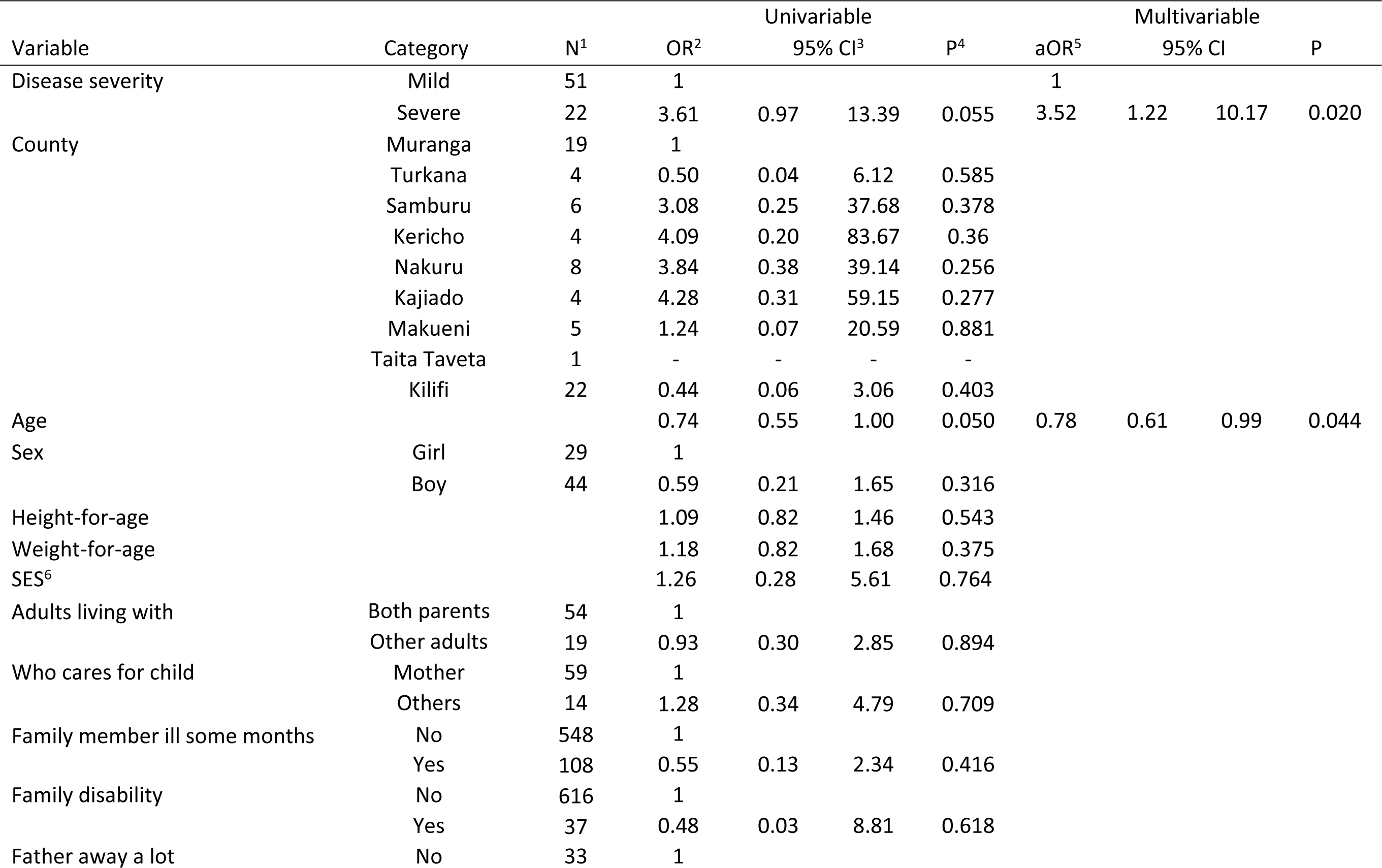
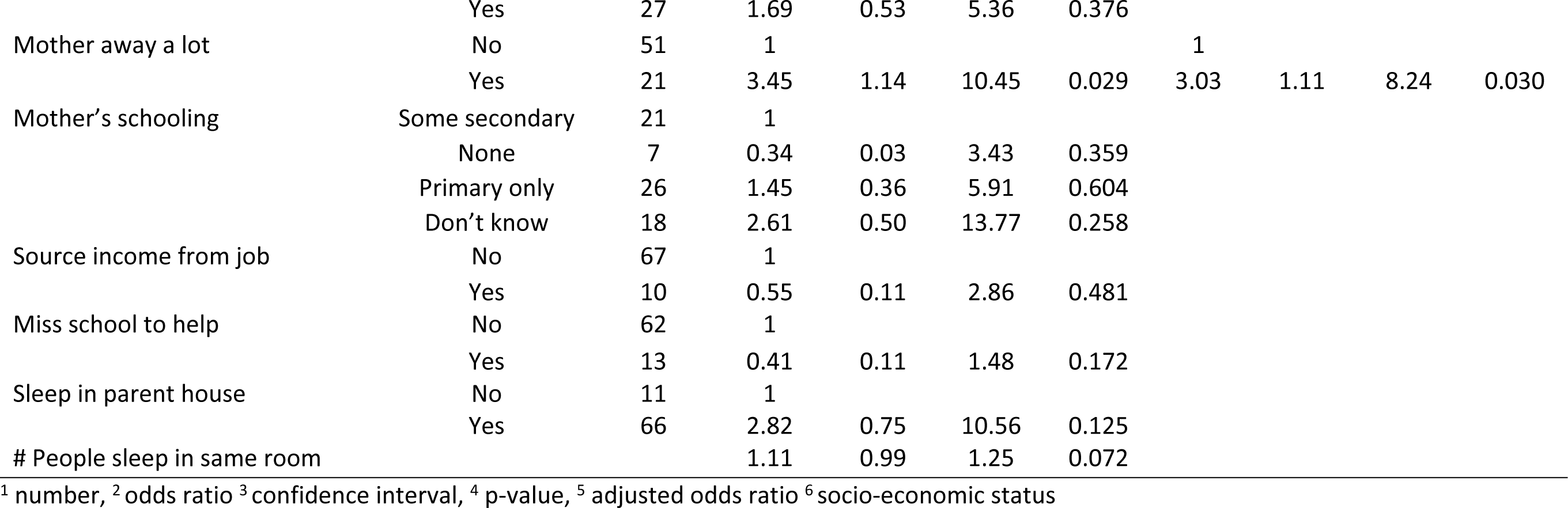
Panel Ordered Logistic Regression analysis for tungiasis-associated Quality of Life (TmDLQI) quintiles (categories) for tungiasis patients.

| Variable | Category | N<sup>1</sup> | OR<sup>2</sup> | Univariable |  |  | aOR<sup>5</sup> | Multivariable |  | P |
| --- | --- | --- | --- | --- | --- | --- | --- | --- | --- | --- |
|  |  |  |  | 95% CI<sup>3</sup> |  | P<sup>4</sup> |  | 95% CI |  |  |
| Disease severity | Mild | 51 | 1 |  |  |  | 1 |  |  |  |
|  | Severe | 22 | 3.61 | 0.97 | 13.39 | 0.055 | 3.52 | 1.22 | 10.17 | 0.020 |
| County | Muranga | 19 | 1 |  |  |  |  |  |  |  |
|  | Turkana | 4 | 0.50 | 0.04 | 6.12 | 0.585 |  |  |  |  |
|  | Samburu | 6 | 3.08 | 0.25 | 37.68 | 0.378 |  |  |  |  |
|  | Kericho | 4 | 4.09 | 0.20 | 83.67 | 0.36 |  |  |  |  |
|  | Nakuru | 8 | 3.84 | 0.38 | 39.14 | 0.256 |  |  |  |  |
|  | Kajiado | 4 | 4.28 | 0.31 | 59.15 | 0.277 |  |  |  |  |
|  | Makueni | 5 | 1.24 | 0.07 | 20.59 | 0.881 |  |  |  |  |
|  | Taita Taveta | 1 | - | - | - | - |  |  |  |  |
|  | Kilifi | 22 | 0.44 | 0.06 | 3.06 | 0.403 |  |  |  |  |
| Age |  |  | 0.74 | 0.55 | 1.00 | 0.050 | 0.78 | 0.61 | 0.99 | 0.044 |
| Sex | Girl | 29 | 1 |  |  |  |  |  |  |  |
|  | Boy | 44 | 0.59 | 0.21 | 1.65 | 0.316 |  |  |  |  |
| Height-for-age |  |  | 1.09 | 0.82 | 1.46 | 0.543 |  |  |  |  |
| Weight-for-age |  |  | 1.18 | 0.82 | 1.68 | 0.375 |  |  |  |  |
| SES<sup>6</sup> |  |  | 1.26 | 0.28 | 5.61 | 0.764 |  |  |  |  |
| Adults living with | Both parents | 54 | 1 |  |  |  |  |  |  |  |
|  | Other adults | 19 | 0.93 | 0.30 | 2.85 | 0.894 |  |  |  |  |
| Who cares for child | Mother | 59 | 1 |  |  |  |  |  |  |  |
|  | Others | 14 | 1.28 | 0.34 | 4.79 | 0.709 |  |  |  |  |
| Family member ill some months | No | 548 | 1 |  |  |  |  |  |  |  |
|  | Yes | 108 | 0.55 | 0.13 | 2.34 | 0.416 |  |  |  |  |
| Family disability | No | 616 | 1 |  |  |  |  |  |  |  |
|  | Yes | 37 | 0.48 | 0.03 | 8.81 | 0.618 |  |  |  |  |
| Father away a lot | No | 33 | 1 |  |  |  |  |  |  |  |
|  | Yes | 27 | 1.69 | 0.53 | 5.36 | 0.376 |  |  |  |  |
| Mother away a lot | No | 51 | 1 |  |  |  | 1 |  |  |  |
|  | Yes | 21 | 3.45 | 1.14 | 10.45 | 0.029 | 3.03 | 1.11 | 8.24 | 0.030 |
| Mother's schooling | Some secondary | 21 | 1 |  |  |  |  |  |  |  |
|  | None | 7 | 0.34 | 0.03 | 3.43 | 0.359 |  |  |  |  |
|  | Primary only | 26 | 1.45 | 0.36 | 5.91 | 0.604 |  |  |  |  |
|  | Don't know | 18 | 2.61 | 0.50 | 13.77 | 0.258 |  |  |  |  |
| Source income from job | No | 67 | 1 |  |  |  |  |  |  |  |
|  | Yes | 10 | 0.55 | 0.11 | 2.86 | 0.481 |  |  |  |  |
| Miss school to help | No | 62 | 1 |  |  |  |  |  |  |  |
|  | Yes | 13 | 0.41 | 0.11 | 1.48 | 0.172 |  |  |  |  |
| Sleep in parent house | No | 11 | 1 |  |  |  |  |  |  |  |
|  | Yes | 66 | 2.82 | 0.75 | 10.56 | 0.125 |  |  |  |  |
| # People sleep in same room |  |  | 1.11 | 0.99 | 1.25 | 0.072 |  |  |  |  |

<sup>1</sup> number, <sup>2</sup> odds ratio <sup>3</sup> confidence interval, <sup>4</sup> p-value, <sup>5</sup> adjusted odds ratio <sup>6</sup> socio-economic status

## Discussion

Few of the countries endemic for tungiasis have a national control strategy. This is in part due to lack of data on the disease burden and evidence-based interventions, but also a lack of appreciation for the impact the disease has on those who are infected. Here we attempted to quantify the impact with the aim of generating evidence to advocate for the disease to be put on local, national and international agendas.

The study found that tungiasis infection is negatively associated with weight-for-age, with higher levels of school absenteeism and lower exam scores among pupils in the younger classes. Children with severe tungiasis had higher levels of pain and itching, and a reduced quality of life, particularly as regards mobility, concentration in school and disturbed sleep, than children with mild tungiasis.

The most striking impact identified in this study was the very poor academic achievement of infected pupils in grade one to four in all three exam subjects compared to uninfected peers, even when adjusted for other factors that can influence school performance, including absenteeism which was independently associated with tungiasis. This aligns with what has been reported for other diseases. Previously children with repeated episodes of malaria[21, 22] or with helminth infections [23, 24] have been found to have lower school attendance, learning ability, memory and academic achievement. It is quite possible that our result could have been confounded by the children in our study being co-infected with these other diseases since we did not test for them. While the comparison group of uninfected pupils may have controlled for this, it is possible the diseases cluster in the same children. Future studies on academic impact should test pupils for malaria and helminth infections as well as tungiasis.

Another factor that could be a confounder in the association with school performance is malnutrition, low height-for-age (stunting) and weight-for-age (underweight), both of which have previously been associated with poor academic achievement [25–27]. However, in the current study only height-for-age was positively associated with math scores in the lower grades in this study and did not confound the association with tungiasis, so we find it likely that tungiasis independently affects children’s school performance.

Stunting is thought to be caused by chronic under-nutrition and is associated with poor neurological and cognitive development and therefore learning ability and academic achievement[28, 29]. It is possible that the chronic pain leads to poor nutritional intake, but also possible that poverty is a causal factor that confounds the association between nutrition and tungiasis. The mechanism of action for tungiasis infection on poor school performance needs to be explored further but could be related to the inflammation induced by the embedded fleas causing extreme pain and itching as demonstrated in this study, particularly children with severe disease. This in turn affects mobility, sleep, concentration in class, and social exclusion as seen in the quality-of-life assessments and ability to attend school, which could all impact cognitive development and learning. In fact findings of ours from another study demonstrated school children with tungiasis did have impaired cognitive development compared to their uninfected peers[30].

All of these factors are interconnected in a vicious cycle as illustrated in Figure 2, possibly trapping children into a life of poverty[2] which puts them at greater risk of tungiasis. The mechanism of association of tungiasis with weight-for-age even when adjusted for SES needs further investigation but may be a result of this cascade. The embedded fleas and the associated itching and mental state may affect a child’s appetite, or the fleas could be depriving the host of essential nutrients, particularly those with a high infection intensity.

**Figure 2.**
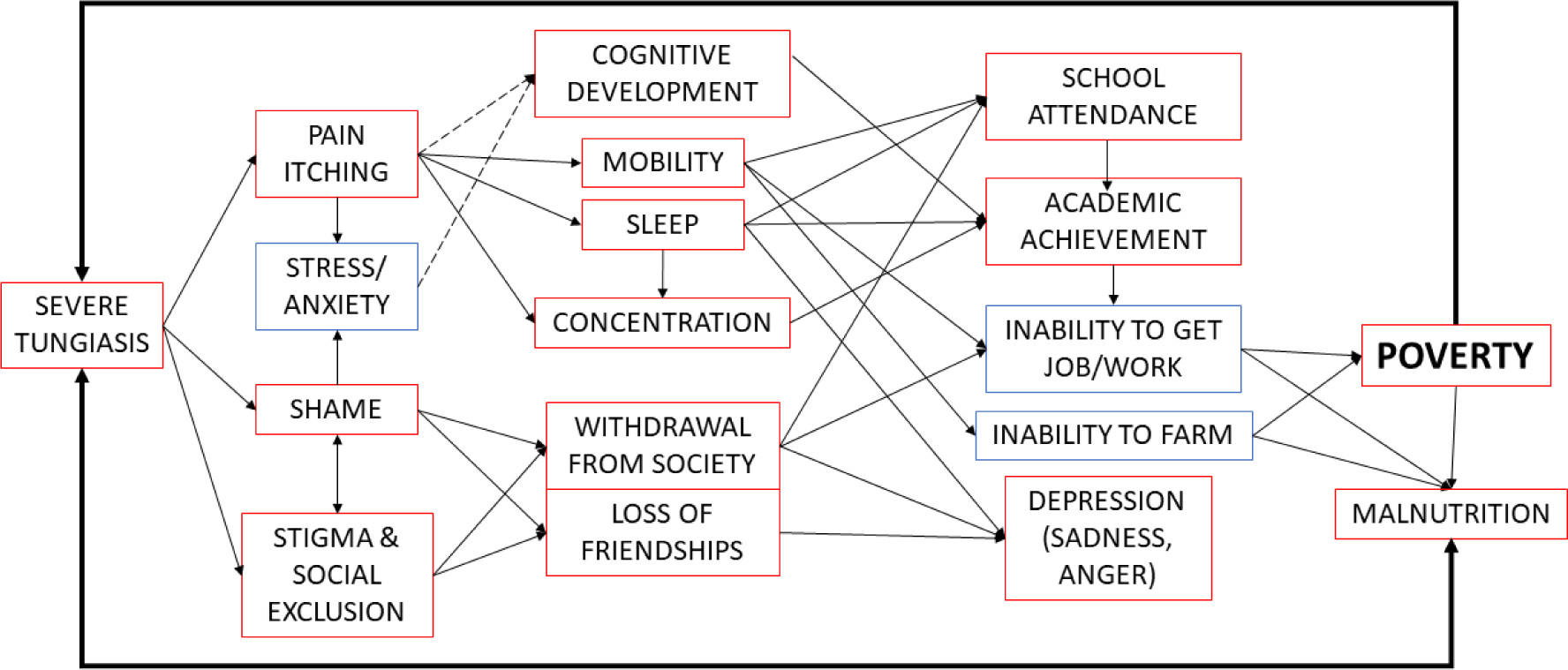
The network of impacts of tungiasis trapping people in poverty. Red boxes documented impacts, blue boxes anecdotal reports, solid arrows: likely pathway, broken arrow: possible pathway.

The proportion of infected pupils (68.5%) who reported a moderate to very high impact on their TmDLQI was similar to the previous small tungiasis study in Kenya (78%)[13] and a study of cutaneous larva migrans in Brazil (71%) [31], which were higher than that reported for scabies (45%) [32], suggesting tungiasis has a higher impact on the quality of life of children than does scabies, another skin disease caused by an arthropod. Each of these studies used a disease-specific, modified version of the general CDLQI[17]. Both Kenyan tungiasis studies identified sleep disturbance and concentration in school as the most affected domains which have been shown to lead to poor cognitive development[33], learning and memory[34] as well as depression[35].

We have recently proposed a new classification of disease severity for tungiasis using a 10-flea threshold. The fact that children with severe tungiasis in this randomly selected population from multiple geographic areas and cultures across Kenya expressed five times higher levels of pain and itching and a higher impact on quality of life, mobility and concentration confirms this is a clinically relevant classification for future use in other studies, and for targeting and monitoring interventions.

There were several possible limitations to this study. One limitation of the academic outcome is the possibility that the scoring system for this age group by teachers could be subjective and biased. However, the schools were selected randomly and were from nine different counties across Kenya and it is unlikely all the teachers would have the same bias. Using the school as a random effect in the models should also control for this. If this association reflects a negative bias of teachers against pupils with tungiasis rather than an actual difference in academic achievement, that would also be an important outcome and barrier to these children’s development which would require interventions. Further studies to investigate this might be warranted.

Another limitation is that participants were selected from children who were attending school on the day of the surveys. The most heavily infected children in the community may have been absent from school on that day on account of the disease and may miss more days of school due to their infection. If we had enrolled such children, we would expect to have seen even higher impacts on absenteeism rates and even lower exam scores.

The lack of association of tungiasis with exam scores in the older age group (grade 5 to 8) may well be the result of the very small number of cases enrolled in the study. Ideally this study should be repeated in public schools only and enrolling an equal number of cases and controls, across all school grades into a longitudinal study to determine whether older pupils also have lower exam scores and whether the impact seen at the younger age affects achievement in later years.

Lastly, although we examined the pupils for other skin diseases, we did not test them for diseases such as malaria and intestinal helminths which could have confounded several of our outcomes including absenteeism, school grade delay and school performance. However, the considerably larger number of controls and their random selection from randomly selected schools across the nine counties in the study should have gone some way to counter this.

In conclusion, our study has demonstrated that tungiasis has considerable impact on many aspects of the lives and development of children, just as has been documented for malaria and helminth infections. Diagnosis, surveillance and disease management for children with tungiasis urgently needs to be integrated into the existing platforms created for these other diseases, in line with the WHO Road map for Neglected Tropical Diseases[36]. Only by addressing all of the diseases impacting these marginalized children will they have an equal opportunity to develop alongside their uninfected peers.

### Ethics approval and consent to participate

The study was approved by the KEMRI Scientific and Ethics Review Committee (approval number KEMRI/SERU/CGMR-C/170/3895) as well as the Oxford Tropical Research Ethics Committee (reference number 38-19). The study was conducted in accordance with the Helsinki Declaration. During the community engagement phase, a presentation was made to the national Director for Neglected Tropical Diseases at the Ministry of Health, the county and sub-county health management teams and the department of education in all counties to obtain their approval. In each school a meeting was held with the school parent teachers’ association (PTA) or management board to obtain their permission to conduct the survey in their school. The head teacher and PTA chairperson signed the consent form on behalf of the parents and school for the pupils to be examined. Each child gave verbal assent. Community health workers were hired and trained in each school to assist and be the link with the community emphasizing that participation was completely voluntary, and subjects had the opportunity to withdraw from the study at any point in the study.

For pupils selected for interviews, these were explained, and each was given an information leaflet to take home for their parents along with an opt-out form. Parents were to sign and return the form only if they did not want their child to participate in the interviews, or they could attend the school the next day to clarify any issues they may have. On the following day, if the selected pupils did not have the opt out form and were willing to participate, they proceeded with the interviews.

All data were collected on PIN protected electronic tablets, stored on password protected RedCap databases on the KEMRI-Wellcome Trust servers. Data were analyzed after export to Excel spreadsheets without inclusion of personal identifiers.

All pupils with tungiasis were referred for treatment to the community health workers or the local health facility using benzyl benzoate provided by the study. For those with secondary bacterial infection and other illnesses requiring treatment, a referral was made to the nearest health facility.

### Consent for publication

Not applicable

## Data Availability

The datasets and other materials supporting the conclusions of this article are available on KWTRP Research Data Repository at Harvard Dataverse through the following link: https://doi.org/10.7910/DVN/DFSTIZ

Elson Tungiasis Impact dataset

## Supporting Information

SI 1. Table of participant distribution by covariates and disease status

SI 2. Univariable Regression tables for pupil exam results in Maths, English and Science

SI 3. Frequency histogram of TmDLQI with quintile thresholds.

## Author Contributions

Conceptualization, LE, UF; Methodology, LE, UF, BO; Formal Analysis, LE, BO; Investigation, LE, CK, SK, CM, MO, ES, ES, JK, EM, MW, JL; Resources, LE, PB; Data Curation, LE ; Writing – Original Draft Preparation, LE.; Writing – Review & Editing, UF,BO, PB; Visualization, LE.; Supervision, PB; Project Administration, LE, PB; Funding Acquisition, LE, PB.

## Acknowledgements

We are grateful to the communities who participated, the school Parent Teacher Associations and Head Teachers who allowed us to work in their schools, the county Directors of Health and Education who gave their approval for the study and assisted in many ways to ensure its success.

## Abbreviations

aOR: adjusted Odds Ratio
aIRR: adjusted incidence rate ratio
CDLQI: Children’s dermatological life quality index
CI: Confidence Interval
IQR: inter-quartile range
KEMRI: Kenya Medical Research Institute
NTDs: Neglected Tropical Diseases
OR: Odds ratio
SES: Socio-economic status
TmDLQI: Tungiasis modified life quality index
WHO: World Health Organisation

